# Municipal wastewater surveillance reveals socioeconomic and immigration gradients in antimicrobial resistance across Alberta, Canada

**DOI:** 10.64898/2026.07.19.26358431

**Authors:** Jangwoo Lee, Catalina Gonzalez, Emily Au, Nicole Acosta, Barbara J. Waddell, Zhaohui S. Xu, Rhonda G. Clark, R. Benson Weyant, Bruce Dalton, Rahat Zaheer, Tim A. McAllister, Herman Barkema, Diego Nobrega, Srijak Bhatnagar, Bonita E. Lee, Xiaoli Pang, Christine O’Grady, Kevin Frankowski, Stefania Bertazzon, John M. Conly, Casey R.J. Hubert, Michael D. Parkins

**Author notes:** Department of Biology, McMaster University, Hamilton, Ontario, Canada (Jangwoo Lee). Corresponding Author:: Michael D. Parkins, MD, MSc, Professor of Medicine and Microbiology, Immunology & Infectious Diseases, Cumming School of Medicine, University of Calgary, 3330 Hospital Drive NW, Calgary, AB, Canada, T2N 4N1.

## Abstract

Antimicrobial resistance (AMR) is an ever-increasing threat to population health. Industrial, environmental and societal factors are increasingly recognized as important contributors to AMR within communities. Here, we investigated the spatial distribution of AMR genes (ARGs) across Alberta, Canada and their association with socio-economic, immigration-related, and agro-industrial characteristics using municipal wastewater-based surveillance. We analyzed monthly wastewater metagenomes collected between March 2022 and March 2023 across eleven municipalities, representing 39% of Alberta’s population. Integration with census data enabled multivariate analysis, revealing that municipal resistome profiles were strongly structured along income and immigration-related population gradients. ARGs spanning 14 resistance classes exhibited distinct distributional patterns across income and immigration gradients, including contrasting associations among beta-lactam, aminoglycoside, and macrolide-lincosamide-streptogramin ARGs, consistent with heterogeneous selection pressures across sub-populations. These findings demonstrate the capacity of longitudinal wastewater surveillance to identify persistent population-level resistome patterns and highlight the importance of incorporating sociodemographic context into AMR surveillance and mitigation strategies.

**Highlights:**

- Monthly wastewater surveillance captured year-round provincial resistome patterns
- Multivariate analyses disentangled societal associations with resistomes
- Income and immigration gradients were strongly associated with resistomes
- Wastewater surveillance supports equitable population-level AMR monitoring

## Introduction

Antimicrobial resistance (AMR) is an escalating threat to population health. Globally, 1.27 million deaths were attributable to AMR in 2019, and this toll is expected to increase to ≥8.5 million by 2050 (Murray et al., 2022; Naghavi et al., 2024). In addition to health outcomes, AMR has significant economic consequences. In Canada alone, hospital-associated direct costs reached USD$1.4 billion in 2018, and are projected to rise to as much as USD$6 billion by 2050 (Diener et al., 2022).

Antimicrobial use is a primary driver of AMR, with higher antimicrobial consumption consistently associated with increased resistance across diverse pathogens and geographic regions. For example, data for the year 2020 revealed significant correlations between beta-lactam and fluoroquinolone consumption and resistance in clinically isolated *Escherichia coli* and *Klebsiella pneumoniae* spanning 14 different European, Asian and African countries (Ajulo and Awosile, 2024). These relationships highlight the central role of human antimicrobial use in shaping population-level resistance patterns. Antimicrobial use, however, extends far beyond the human sector.

The drivers of AMR, however, extend beyond human antimicrobial use. The livestock industry is one of the most intensive antibiotic users in Canada; the quantity of antibiotics sold for animals was up to 4.1 times greater than the quantity used in humans, including for several of the antibiotic classes most commonly used in human medicine between 2018 and 2021 (Government-of-Canada, 2023), underscoring its importance as a reservoir for the selection and amplification of AMR. Within the broader One Health context, interactions between human, animal, and environmental reservoirs contribute to the complexity of AMR transmission dynamics. Individuals working with livestock may therefore represent a group disproportionately exposed to AMR. More broadly, up to 60% of human pathogens are estimated to originate from animals (Ikhimiukor et al., 2022), and evidence demonstrates zoonotic acquisition of AMR (Knetsch et al., 2018; Lupindu et al., 2015).

More recently, antimicrobial prescribing has been shown to vary across sub-populations as a function of multiple societal variables. For instance, a study performed in the United States found that children from lower socio-economic status (SES) communities tended to receive more antibiotics, associated with a higher risk of AMR development (McGurn et al., 2021). A structured narrative review conducted in Canada on the relationship between SES and AMR found that higher income was generally negatively associated with community-acquired methicillin-resistant *Staphylococcus aureus* (King et al., 2022). Immigration and international mobility represent additional important determinants of AMR. Given well-documented global variation in antimicrobial resistance, population movement between regions may contribute to changing local resistance patterns through the introduction and dissemination of resistant organisms. Many studies have similarly demonstrated increased carriage of resistant organisms among individuals with recent international travel, healthcare exposure or immigration from regions with elevated AMR prevalence, suggesting that global interconnectedness may contribute to local resistome composition (Bacaner et al., 2004; D’Souza et al., 2021; dos Santos Tufic-Garutti et al., 2021). In Canada, the income gap between foreign-born and Canadian-born residents has been substantial and has continued to increase since 1980 (Crossman, 2013). The economic disparity between foreign-born and Canadian-born residents can impact individuals’ opportunities across various social domains, as evidenced by associations between immigration status and multiple socioeconomic indicators (StatsCan, 2022b). Collectively, these findings underscore the interconnection between SES, immigration and AMR within the Canadian context, highlighting the need for integrated analyses examining these variables together.

At a population level, it has become evident that antimicrobial use and exposure are not uniformly distributed, but are shaped by underlying societal and demographic factors. There have been several studies exploring association between AMR and various societal factors (i.e., SES, immigration, and agro-industrial sectors within populations) (Allel et al., 2023; Kurowski et al., 2021; Wozniak et al., 2022). However, most have employed traditional epidemiological approaches that depend on culturing AMR organisms from a relatively limited number of individuals or animals, followed by genotyping and/or phenotyping (Allel et al., 2023; Kurowski et al., 2021; Wozniak et al., 2022). As such, the complex relationship between AMR and societal factors has not been fully elucidated at a large population scale, highlighting the need for complementary approaches. Wastewater-based surveillance (WBS) is an emerging approach that inclusively and comprehensively assesses entire populations, enabling AMR to be measured across a range of scales (i.e., buildings or neighborhoods to whole municipalities) and evaluated in the context of local socio-demographic characteristics (Gupta et al., 2024; Lee et al., 2025). Recently, our group demonstrated at the city level that associations between population-level AMR and societal factors are dynamic and may depend on the scale of analysis (Lee et al., 2025). A nationwide study in the United States investigated the distribution of selected ARG markers by ddPCR and examined their associations with various societal variables based on samples collected during a one-week period in May 2024 (Kim et al., 2026). Together, these studies suggest that societal factors are associated with population-level AMR, but the nature, consistency, and temporal stability of these relationships remain incompletely understood across broader geographic regions. To improve the characterization of population-level ARG profiles and the representativeness of AMR surveillance, WBS should be complemented by longitudinal sampling, metagenomic sequencing, and robust multivariate statistical approaches. Together, these methods capture year-round resistome dynamics and enable the analysis of highly multidimensional datasets.

Here we examined the geospatial distribution of ARGs across the province of Alberta (population 4.3 million), Canada, using WBS to test the hypothesis that societal and agro-industrial factors influence their distribution in human populations. We further hypothesized that a provincial-scale analysis would provide sufficient socio-demographic and industrial heterogeneity to identify meaningful associations with population-level resistome composition, while reducing some of the local variability and international confounding that can complicate analyses conducted at smaller or larger geographic scales. Alberta provides an informative setting in which to evaluate this hypothesis because healthcare is delivered through a single unified provincial health system (AHS, 2023). The province is home to large and diverse immigrant populations originating from a broad range of countries and socio-economic contexts, creating substantial variation in immigration-related demographic characteristics across municipalities (StatsCan, 2021). Alberta also has the highest income inequality among Canadian provinces (Statistics-Canada, 2023) and contains one of Canada’s largest cattle industries (StatsCan, 2017), providing marked variation in several factors hypothesized to influence population-level resistome composition. By serially monitoring community resistomes over 13 months, we captured year-round ARG dynamics and used multivariate analyses to disentangle their associations with societal variables. The objectives of this study were therefore to (1) examine the distribution of ARGs across the province; (2) identify key societal factors associated with their distribution; and (3) determine whether specific ARGs correlate with individual societal and agro-industrial factors.

## Methods

### Setting, site descriptions and field sampling

We obtained serial raw sewage samples from 11 municipal wastewater treatment plants (WWTPs) across Alberta, representing 39% of the total population of Alberta, Canada (Fig. S1 and Supplementary-Data-S1). Each WWTP served populations ranging from 6,935 to 1,291,770 individuals. Details of population characteristics, socio-economic status and associated agriculture/agrifood industry composition are available in Supplementary Data S1. Autosamplers (ISCO5800 or ISCO6712, Teledyne ISCO, USA) collected a 24-h composite sample once per month from March 2022 – March 2023 (a total of 13 months; see Supplementary-Data-S2), collecting 100 mL every 15 min. Composite samples were shipped on ice, and maintained at 4°C, and processed within 48 h.

### Sample processing and DNA extraction

After thorough mixing, raw wastewater was aliquoted into 120 mL sterile conical tubes, centrifuged (4,000xg at 4°C for 20 min) and the supernatant was removed. The pellets were transferred into sterile 2 mL microcentrifuge tubes and stored at –80°C until further analysis. Following overnight thawing at 4°C, genomic DNA was extracted from 250 μL of the resuspended wastewater pellets using the PowerSoil DNA Isolation Kit (Qiagen, Germany) following the manufacturer’s protocol. DNA concentration and quality (i.e., 260/280 and 260/230 absorbance ratios) were assessed using a NanoDrop spectrophotometer (Thermo Fisher Scientific, USA), and concentrations were confirmed using the Qubit™ dsDNA High-Sensitivity Kit (Thermo Fisher Scientific, USA). An extraction blank was included in every batch, and ensured there was no contamination throughout the protocol (<0.1ng/μL, measured using Qubit High-Sensitivity Kit). The extracted samples were stored at -80°C until further analysis.

### Sequencing and bioinformatic analysis

Frozen extracted DNA was sent to Novogene Corporation Inc., California, USA (https://www.novogene.com/us-en/) for sequencing. Libraries were prepared and shotgun sequencing was performed using Illumina NovaSeq 6000 platform yielding paired-end 2x150bp reads. The final sequencing output was an average of 11.3 gigabases (see Supplementary Data S2). Raw reads underwent quality control first at Novogene by removing reads with N>10% and Qscore≤5, and those containing adapters. Quality was further assessed using FastQC (v0.11.9) (Andrews, 2010), and the reads were further filtered using Trimmomatic (v0.39) (Bolger et al., 2014), when necessary. ARGs were annotated from filtered reads using the DeepARG-SL pipeline v1.0.2 (Arango-Argoty et al., 2018), following established protocols (Acosta et al., 2025; Arango-Argoty et al., 2018; Lee et al., 2025). The pipeline merges paired-end reads, annotates ARGs against its own database compiled from multiple comprehensive open-access databases (i.e., CARD, ARDB, and UniProt for ARGs, and Greengenes for 16S rRNA genes) using a local aligner, and then further screens ARG-like reads through deep-learning algorithms, thereby minimizing false-positive annotations (Arango-Argoty et al., 2018). The final outputs of this pipeline were further processed using a previously published custom script, generating ARGs per 16S rRNA gene as the final metric (Acosta et al., 2025).

We compared our data with 46 published metagenomic datasets from Southern Alberta to better understand the potential influence of cattle density (NCBI-SRA BioProject PRJNA420682) (Zaheer et al., 2019). The samples were categorized as composite feedlot cattle feces (27), runoff samples from the feedlot (13), and raw sewage samples obtained from the WWTPs, including one from our study site (i.e., Calgary) (6) (Zaheer et al., 2019). For bioinformatic analysis, we followed the same procedures described above for wastewater. In brief, the raw reads were quality controlled using FastQC and Trimmomatic, followed by ARG annotation utilizing the DeepARG-SL pipeline v1.0.2 and calculation of ARG abundance metrics. The original study reported that tetracycline resistance genes – the most prevalent ARGs in cattle-related sources, occurred most abundantly in feedlot feces, followed by feedlot runoff and domestic sewage (Zaheer et al., 2019). The levels of ARGs identified in this study were similar to those observed in our dataset (Fig. S2). Overall, these findings confirmed that our workflows were well aligned.

### Analysis of societal variables

Raw data on key socio-economic variables (income, education, and unemployment), and industrial activities (agriculture and agrifood) for each municipality were obtained from the open-access provincial census databases (Government-of-Alberta, 2025). We used data for the most recent year available for each parameter (i.e., 2020 or 2021, depending on the data). We used median values for family income (in $CAD, based on 2020 data), unemployment rate, and three livestock parameters, representing farming activities across the province (i.e., the density (in the number per 1,000 inhabitants) of farms, pigs and cattle in 2021) as income, unemployment, and livestock industry parameters respectively, and these data were directly available from Alberta census data (Government-of-Alberta, 2025). To measure education, we divided the number of people with post-secondary education degrees by the total population in each municipal sewershed available through the provincial open-access portal (Government-of-Alberta, 2025).

To understand the population characteristics, we assessed the proportion of each community composed of first-generation Canadians. For these immigration parameters, we calculated the following four variables: the proportion of first generation Canadians whose places of birth were in countries with high HDI, upper-middle HDI, lower-middle HDI and low-HDI as defined by the United Nations Development Programme (UNDP, 2022a; b). These variables were calculated using parameters sourced from Canadian federal census databases (StatsCan, 2021), as follows : (1) number of immigrants (in 2021) normalized by total population in each city; (2) number of immigrants from each country of birth divided by the total number of immigrants for each city; (3) the formerly calculated two values (from (1) and (2)) were multiplied to calculate ‘the proportion of immigrants from each place of birth relative to the total population’ in each municipality; and (4) the values calculated for (3) were aggregated by HDI category – low for Q1 (the lowest 25%), lower-middle for the Q2 (<50%), upper-middle for the Q3 (50 – 75%), and high for Q4 (the top 25%) (see Fig. S3a), following the rationale outlined in our previous study (Lee et al., 2025). As a result, the proportions of birth countries, categorized into four groups (i.e., high-HDI, upper middle-HDI, lower-middle-HDI, and low-HDI) among first-generation Canadians were calculated for each city or community included in this study (Fig. S3b).

### Statistical analyses

We performed a multivariate analysis between human wastewater ARGs and key societal factors derived from detailed federal census data (i.e., SES, immigration, and livestock industry variables such as farm, cattle, and pig density), using Canonical Correspondence Analysis (CCA) in R using the Vegan (v2.6-4) package (Oksanen et al., 2022), to identify potential contributors to differential spatial distribution of wastewater ARGs across sites. Multivariate analysis, including CCA has its advantages over multiple testing using univariate methods, including minimization of type 1 error (false-positive), and informing on correlation strengths and directions across variables in a single analysis (Meloun and Militky, 2011; Rudy et al., 1992). Independent variables were selected using forward stepwise modelling. The ‘minimum adequate model’ was built using the function ‘ordistep’ in Vegan (v2.6-4), following the methods detailed by Oksanen (2025) (Oksanen et al., 2022). For each forward selection run, the model predictability assessed by ‘Akaike’s Information Criterion (AIC)’ was calculated, after each variable was added, starting from a null model (i.e., ARG ∼ 1). As a result, the model with the lowest AIC, which was considered to exhibit the highest predictability was selected, and it was turned out to be the model including every societal variable (the final AIC = -22.62). The model p-value was calculated using the Chi-Squared test (p<0.001), and the significance of each term (i.e., societal variable) was derived using Permutation Test (n=1000, and p≤0.066 see Supplementary-Data-3) using Stats (v4.1.2).

The ARG subtypes that had the strongest correlations with the relevant societal variables (i.e., cattle density, and annual household income) from the CCA results were identified according to the following procedure: The residual sum of squares (RSS) was calculated between each societal variable (in red vector) and coordinates for ARGs for each variable, and the 200 ARGs with the lowest RSS were identified (Supplementary-Data-4 & 5). A total of 200 ARGs identified through the aforementioned procedure were further correlated with each societal variable using Spearman correlation with multiple testing correction using the Benjamini–Hochberg method using Stats in R.

For group-comparison, we performed a Kruskal-Wallis test using Stats. To identify pairs exhibiting statistically significant differences, a pairwise Wilcoxon test was performed using Stats in R. p-values were corrected for all multiple comparisons according to the Benjamini-Hochberg method using Stats in R (v4.1.2).

## Results

### Tracking longitudinal trends of wastewater ARG abundances across Alberta

A total of 1159 ARG subtypes corresponding to the 29 major classes were identified from 138 wastewater metagenomes, obtained from 11 cities and towns across Alberta, Canada, representing 39% of the province’s population (Fig. S1 and Supplementary-Data-1), over the course of monitoring (13 months; March 2022 – March 2023, analysed monthly). The sampled communities were geographically and demographically diverse, spanning major urban centres, mid-sized municipalities, and smaller rural communities distributed throughout both northern and southern regions of the province, and exhibiting substantial heterogeneity in socioeconomic characteristics, agricultural activity, and immigration profiles.

Among the ARG classes identified, those conferring resistance to the top five consumed antibiotic classes in the region were beta-lactams followed by fluoroquinolones, macrolide-lincosamide-streptogramin (MLS) agents, sulfonamides, and tetracycline antibiotics (Government-of-Canada, 2023). Notably, both fluoroquinolone and sulfonamide resistance genes exhibited distinct temporal trends for certain sites (Fig. 1a). For example, the relative abundances (i.e., 16S rRNA-normalized ARG abundances) of fluoroquinolone resistance genes in Lethbridge increased sharply during the 3^rd^ quarter (July – August) of 2022, whereas sulfonamide resistance genes in Brooks and Lacombe increased during the second to third quarter (April – August) of 2022. In contrast, the remaining ARG classes (i.e., MLS, beta-lactam, and tetracycline) exhibited relatively stable trends and did not change significantly over the monitoring period (Fig. S4a). Despite transient fluctuations in selected ARG classes, overall resistome composition remained relatively stable over time, with differences between municipalities consistently exceeding month-to-month variation within individual communities (Figs. 1a & S2a).

**Figure 1.**
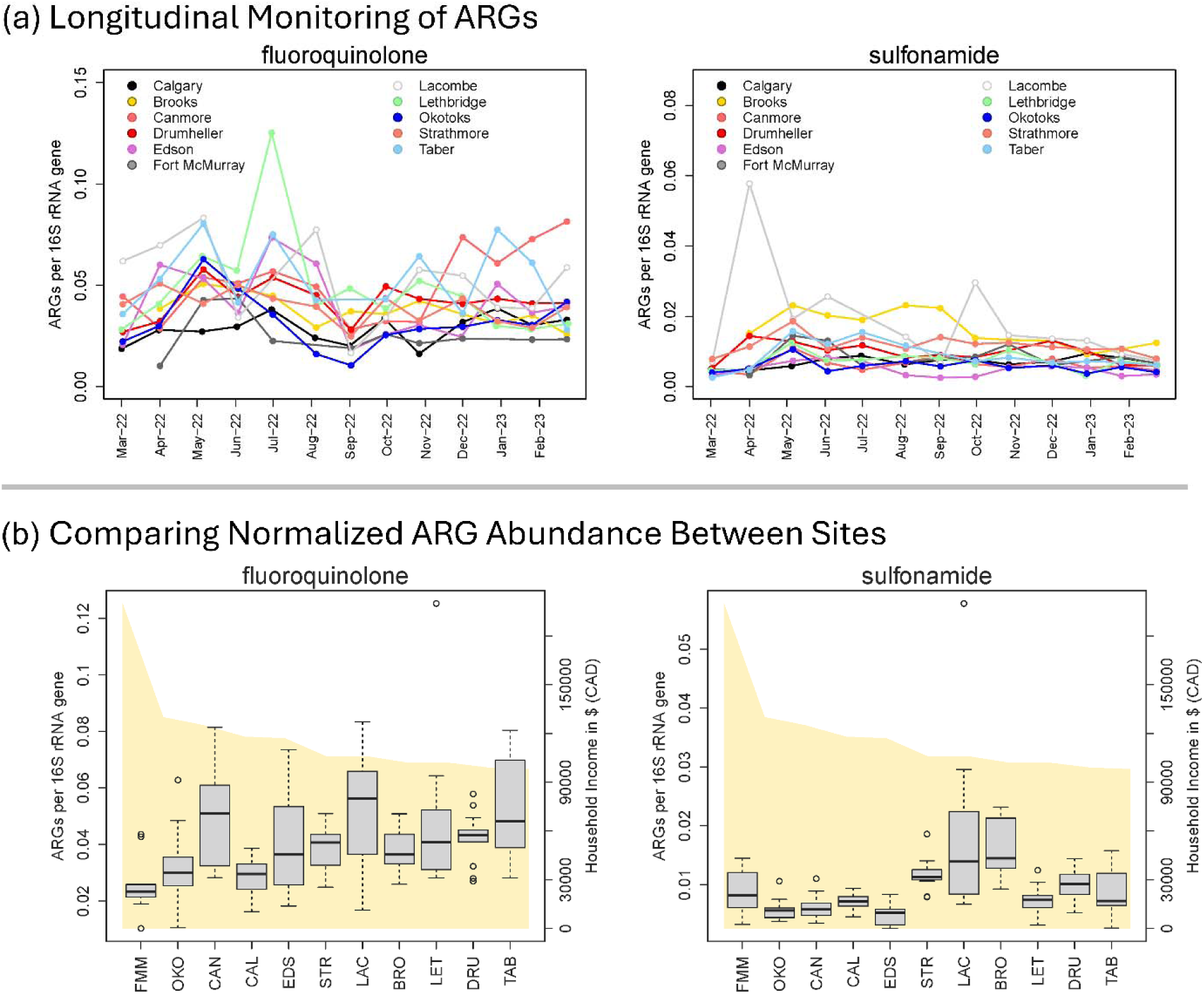
Relative abundances of antimicrobial resistance genes (ARGs) conferring resistance to two of the most frequently used antibiotic classes in Canadian prairies over the course of monitoring (13 months from March 2022 to March 2023), exhibiting visible temporal trends for few sites (i.e., LET, BRO, and LAC). (a) Longitudinal trends of normalized ARG abundances. (b) Comparisons of normalized ARG abundances across locations; the area plot (in yellow) represents annual household income ($CAD), associated with each site. FMM=Fort McMurray, OKO=Okotoks, CAN=Canmore, CAL=Calgary, EDS=Edson, STR=Strathmore, LAC=Lacombe, BRO=Brooks, LET=Lethbridge, DRU=Drumheller, TAB=Taber.

We then compared sites to assess the relative abundance of each ARG class, where we found significant differences (p<0.001) (Fig. 1b). For example, the relative abundances of fluoroquinolone ARGs for Lethbridge were higher than in all the other sites, and the relative abundances of sulfonamide ARGs for both Lacombe and Brooks were similarly higher than at other sites (Fig. 1b). Furthermore, the relative abundances of ARG classes for MLS agents, beta-lactams, and tetracyclines differed by site (p≤0.002) (Fig. S4b).

### Correlating wastewater ARGs with societal factors

To identify factors associated with wastewater ARGs composition across municipalities, we performed multivariate analyses incorporating socio-economic, immigration-related, and livestock-associated variables derived from provincial census and agricultural datasets. According to the forward selection, the model with the lowest Akaike Information Criterion (AIC, ranging from 5.09 to -22.62, with -22.62 for the selected model) was used for downstream analysis, which included ten societal variables (i.e., income, education, unemployment, farm density, cattle and pig density, and immigration; the immigration variable was categorized into four groups based on individuals coming from the countries with high, upper middle, lower middle, and low Human-Development Index (HDI)) (Fig. 2). The p-value for the model was p<0.001, and individual variable contributions had p-values ≤0.066 (Supplementary-Data-3).

**Figure 2.**
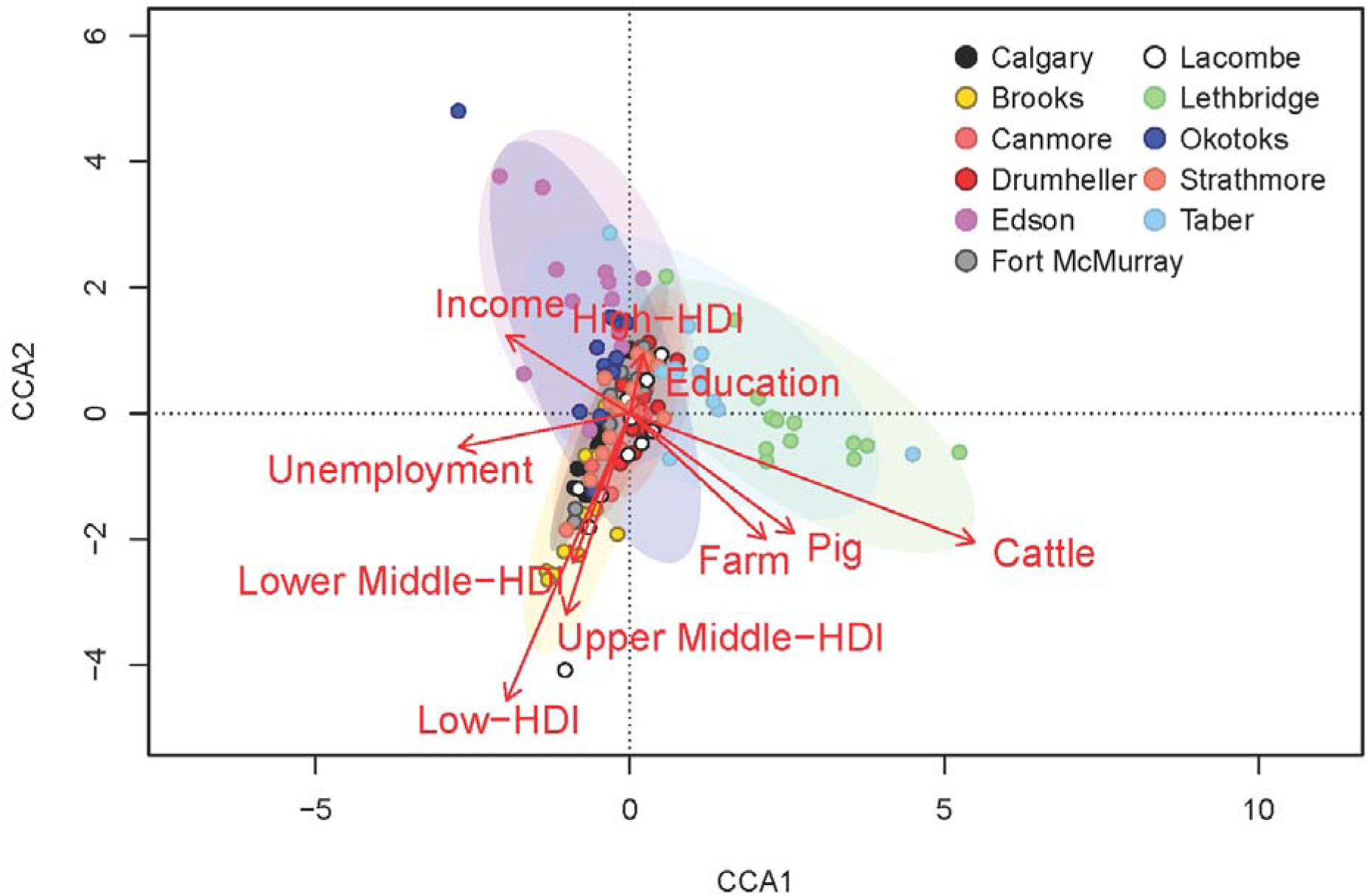
Results of canonical correspondence analysis (CCA), showing associations between ARGs measured in wastewater across Alberta municipalities with multiple societal variables. The strengths and direction of correlation for each societal variable are presented as red vectors. Income represents annual household income ($CAD); education indicates the proportion of individuals who have completed post-secondary education; unemployment represents the proportion of individuals who were not employed; Farm, pig, and cattle denote farm, pig, and cattle density associated with each site. HDI denotes Human Development Index.

Overall, strong associations among specific societal variables were observed, whereas income and unemployment parameters were negatively correlated with livestock-related variables. Education parameters were negatively correlated with multiple immigration parameters, particularly for areas where immigration from low-HDI countries was most prevalent. Income and livestock-related variables were orthogonal to immigration-related variables, indicating that these variables groups were largely independent (Fig. 2). To identify which ARG subtypes were associated with each parameter, we performed biplot analysis, correlating coordinates for ARGs with two of the most significant societal variables, namely cattle density and immigration from low-HDI countries (Fig. S5). We identified the top 200 ARGs most strongly associated with the low-HDI immigration axis, of which 138 genes spanning 15 classes were positively associated, and 62 genes spanning 13 classes that were negatively associated with increasing low-HDI immigration (Supplementary-Data-4). Similarly, for the cattle density axis, 136 ARGs belonging to 17 different classes co-occurred positively with cattle density, whereas 64 genes spanning 16 classes showed negative associations with increasing cattle density (Supplementary-Data-5).

### Resolving competing drivers of ARGs – environment associations

Given that multiple variables were cross-correlated, it was not certain which factors were the primary determinants, particularly for ARGs associated with the cattle density axis; both income and cattle density were moderately correlated with those genes, but in opposite directions (Fig. 2). This finding raised the possibility that some observed associations may be indirect. For example, if livestock-associated occupations are linked to lower income, an apparent association between ARGs and cattle density could arise through underlying socio-economic structure rather than direct transmission (Fig. 3a). Indeed, the agriculture sector, which includes the livestock industry, had the second lowest hourly wage in Canada among 16 industrial sectors (StatsCan, 2022a). Alternatively, both income and cattle density could independently contribute to observed ARG patterns (Fig. 3a). To distinguish between these possibilities, we tested whether ARGs associated with cattle density were enriched in livestock-related sources. Meta-analysis of publicly available metagenomes from Alberta, including feedlot, manure and domestic sewage samples, revealed the opposite pattern to that expected under a livestock-driven model; these genes were more abundant in domestic sewage than manure (p<0.001) and feedlot samples (p<0.001; see Fig. 3b). These findings do not support livestock as the primary determinant and are more consistent with human-associated or socio-economic factors.

**Figure 3.**
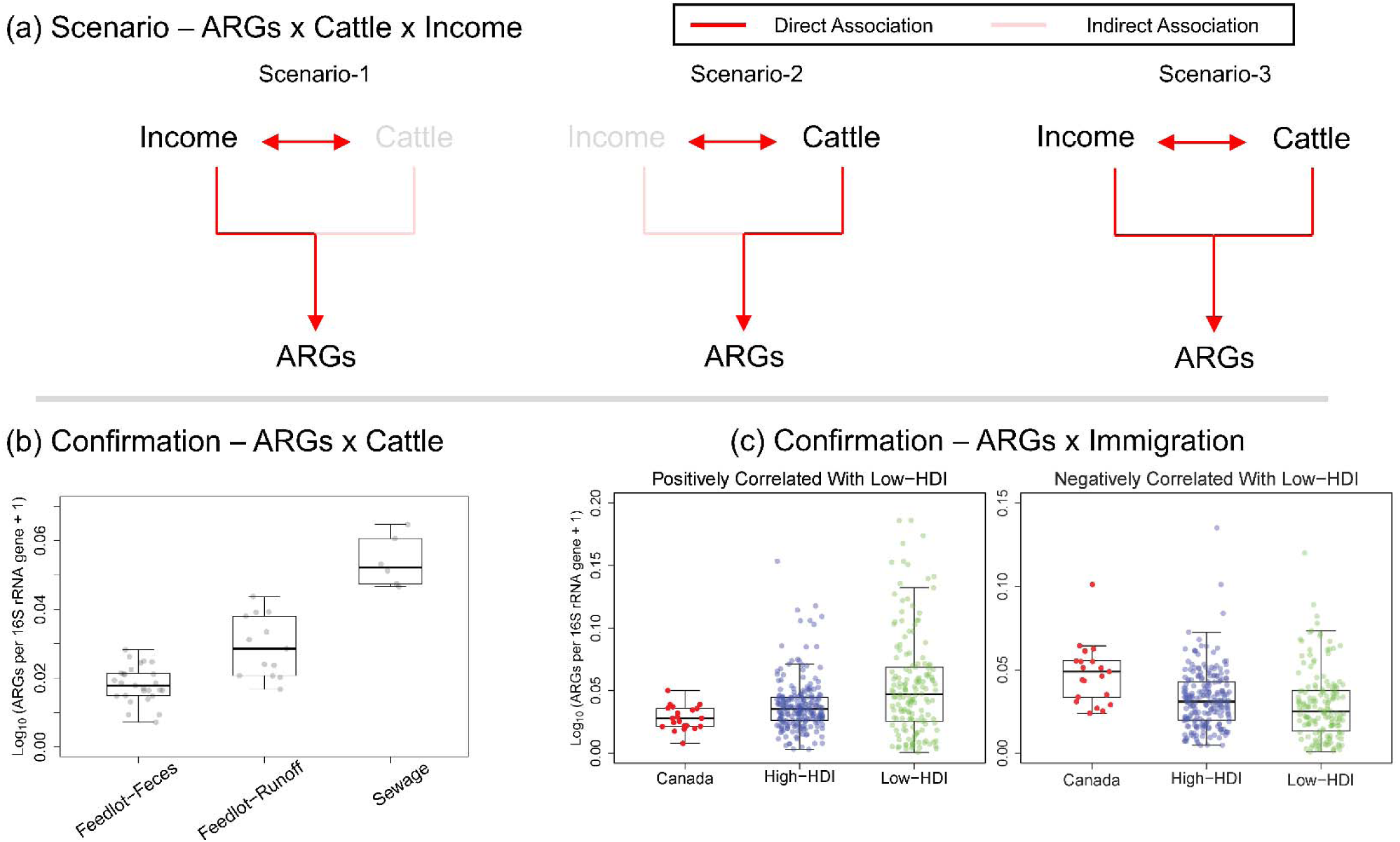
The proposed scenarios explaining how ARGs may be associated with two of the most significant societal variables, identified from Fig. 2 (i.e., income and cattle), and evaluated through meta-analysis. (a) Diagrams visualizing potential relationships among ARGs, income, and cattle; (b) Abundances of ARGs positively correlated with cattle identified through CCA, occurred in cattle manure, feedlot wastewater, and domestic sewage (data sourced from Zaheer et al., 2019); (c) Abundances of ARGs positively associated (left) or negatively associated with low-HDI country immigration (right) identified through CCA, observed across cities in Canada, High-HDI, and Low-HDI countries (data sourced from Hendriksen et al., 2019, and Munk et al., 2021).

We next validated the associations identified in multivariate analysis using univariate Spearman correlation with income and low-HDI immigration. Not all ARGs showed significant associations. A total of 45 out of 200 ARGs spanning 9 classes correlated significantly with low-HDI immigration, among which 31 genes were positively and 14 genes were negatively associated with increasing low-HDI immigration. For income, a total of 109 out of 200 ARGs spanning 13 classes showed significant associations, including 22 positively associated genes and 87 negatively associated genes (Fig. S6 & Supplementary-Data-4 and 5).

When aggregated, positively associated ARG sets increased with low-HDI immigration, whereas negatively associated gene sets decreased (r=0.42 with p<0.001; r=-0.38 with p<0.001, respectively) (Fig. 4a). These relationships were driven primarily by municipalities with the steepest immigration gradients, including Lethbridge, Fort McMurray, Calgary, and Brooks (with low-HDI immigration ranging from 2.6% to 12.0%) (Fig. 4a).

**Figure 4.**
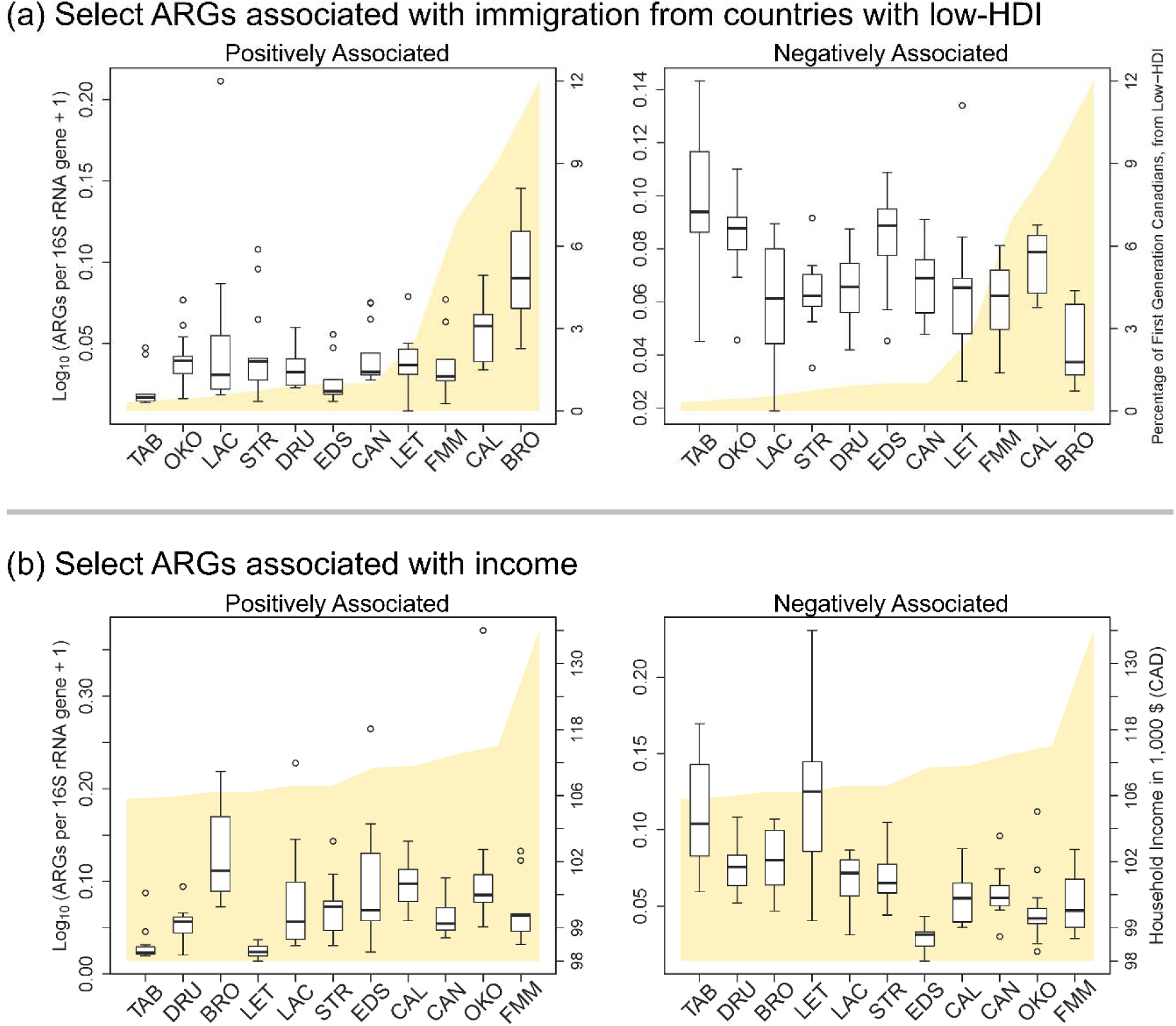
Abundances of key ARGs (aggregated) significantly associated with immigration (i.e., low-HDI) or income parameters, as validated by Spearman correlation. (a) Comparison of relative abundances of key ARGs, positively (left) and negatively associated with low-HDI (right) across sites; (b) Comparison of relative abundances of key ARGs, positively (left) and negatively associated with income (right) across sites.

We wanted to understand the mechanism by which low-HDI immigration was associated with population-level AMR patterns. We hypothesized that ARGs positively associated with low-HDI immigration reflected imported resistome signatures arising through either (1) variations in gut microbiomes and resistomes based on national origin (Deschasaux et al., 2018), or (2) acquisition of AMR through international travel (D’Souza et al., 2021). Accordingly, we expected these genes to be more abundant in settings with greater contributions from low-HDI populations. To test this, we compared the abundances of genes significantly associated with low-HDI immigration (p.adj<0.01) with 656 globally sourced wastewater metagenomes from 244 cities across 101 countries spanning a range of HDI settings (Hendriksen et al., 2019; Lee et al., 2025; Munk et al., 2022). We observed that the abundances of positively correlated ARGs in Canadian wastewater samples (including Alberta samples) were lower than in low-HDI settings (p=0.003) (Fig. 3c); conversely, negatively associated genes were more abundant in Canadian wastewater than in low-HDI settings (p<0.001) (Fig. 3c). These findings support the hypothesis that ARGs positively associated with low-HDI immigration may, in part, reflect external resistome inputs, suggesting that importation contributes to local enrichment across Alberta. Similarly, ARGs associated with income showed consistent directional relationships (r=0.37, p<0.001; r= –0.57, p<0.001) (Fig. 4b). In contrast to low-HDI-associated patterns, income-related associations appeared more gradual across municipalities (Fig. 4b).

The aforementioned ARG sets (positively and negatively associated with low-HDI immigration and income) were also summarized, stratified by major classes (Fig. 5). Among the 16 classes of ARGs identified, multidrug ARGs were the most or second most abundant group overall. Beyond multidrug resistance, distinct resistance-class enrichments were observed, with beta-lactam resistance predominating among ARGs positively associated with low-HDI immigration, MLS resistance among those negatively associated, MLS resistance among genes positively associated with income, and aminoglycoside resistance among those negatively associated with income (Fig. 5).

**Figure 5.**
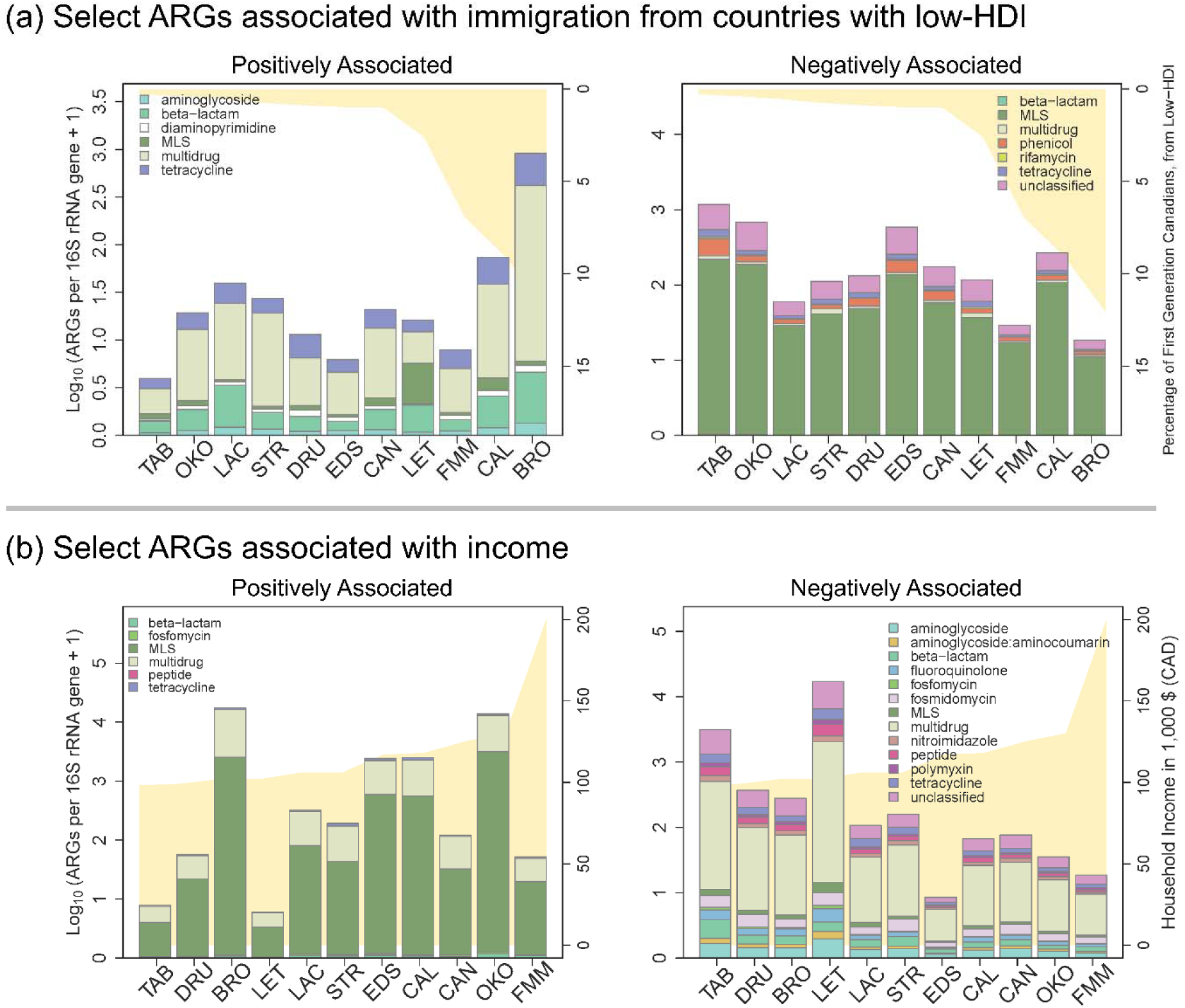
Abundances of key ARGs (by major class) significantly associated with immigration (i.e., low-HDI) or income parameters, as validated by Spearman correlation. (a) Comparison of relative abundances of key ARGs, positively (left) and negatively associated with low-HDI (right) across sites; (b) Comparison of relative abundances of key ARGs, positively (left) and negatively associated with income (right) across sites.

## Discussion

### WBS is a useful tool to understand societal determinants of AMR

Income inequality has increased globally, particularly after the COVID-19 pandemic (Burkinshaw et al., 2022). Given that population health depends on various socio-economic, environmental, and occupational factors, economic inequality leads to health disparities (Group, 2015). Studies have explored the relationship between AMR and societal factors leveraging traditional epidemiological approaches (Allel et al., 2023; Neves et al., 2019). For example, studies performed in Chilean and Brazilian hospitals explored the prevalence of AMR across socio-economically diverse groups utilizing culture-dependent approaches and identified significant associations between societal parameters and AMR in recovered clinical pathogens (Allel et al., 2023; Neves et al., 2019). However, these approaches reflect only a very small number of individuals within an entire population and may not be representative of the entire group. In contrast, WBS has the capacity to analyze the differential distribution of ARG burden across spatial locations and correlate these patterns with the characteristics of populations from which they derive in an inclusive and comprehensive manner. By correlating wastewater-derived ARG burden with census data (Figs. 2 & 4), it is possible to explore associations between AMR and various societal variables. Importantly, the observed associations were identified using monthly metagenomic surveillance conducted over 13 months, suggesting that these relationships reflect relatively stable population-level characteristics rather than transient resistome fluctuations. Recent large-scale wastewater studies have reported similar associations using shorter sampling intervals (Kim et al., 2026); however, longitudinal surveillance may provide additional confidence that observed relationships represent persistent population-level signals.

### Various societal variables are interconnected and influence each other

One of the key observations in our study was that the relationship between ARGs and societal variables was highly complex and dynamic – multiple societal factors were cross-correlated. This implies that relying solely on selected variables in large-scale observational studies may result in misinterpretation of the complex relationships between AMR and its determinants. As an example, a pronounced correlation between ARGs and cattle might not be due to a direct association between them, but rather an indirect effect, caused by cross-correlation between cattle and income, where income might be a determinant (Fig. 3). This indicates that the potential role of human-cattle transmission might not be readily visible in population-level AMR, as it was likely overshadowed by a more dominant factor (i.e., income).

Indeed, income is an important outcome determinant, influencing individuals’ health in multiple ways. For example, income levels are directly related to individuals’ health by limiting access to healthcare services (Lazar and Davenport, 2018). Even though Canada’s publicly funded system has sought to minimize health inequalities by covering outpatient healthcare visits and investigations and all costs related to hospitalizations, lack of universal coverage for outpatient prescriptions continues to be a burden for lower-income Canadians (Bryant et al., 2011). Furthermore, financial strain can also limit individuals’ food choices and eating patterns, potentially serving as a risk factor for various diseases, including common infectious syndromes such as pneumonia, skin and soft tissue infections, and gastroenteritis, which are disproportionately observed in socio-economically disadvantaged populations (Buczkowska et al., 2023; Sood et al., 2023).

Even though our results did not demonstrate a direct association between cattle and AMR at the municipal population level, ARGs can originate from zoonotic sources and be transmitted to humans at a local scale. For example, a study performed in livestock farms housing yaks, sheep, pigs, and horses found that intensive farming significantly altered animal fecal resistomes and also increased phenotypic resistance to many types of antibiotics (Wang et al., 2023). Another study targeting cattle farm workers identified that ARG abundances in the oropharynx of farm workers were significantly higher than those in the general community, and that farm workers’ gut microbiome and resistomes were influenced by farming activities, with variation according to type and duration of work (Ding et al., 2022). While the livestock industry is one of the major drivers of Alberta’s economy, and Lethbridge is an important community in the province’s agricultural sector, workers in agricultural sectors (including the livestock industry) account for only 2.9% of Lethbridge’s labor force (StatsCan, 2021). Given that agriculture sector workers are substantially outnumbered by those in sales and service (26.6%), trade, transport, and equipment operation (18.4%), and business, finance and administration (14.4%) (StatsCan, 2021), the potential influence of cattle-derived ARGs may be diluted at the population-level. To assess agricultural influences on population-level AMR, it may be necessary to undertake more granular surveillance targeting specific sub-populations or smaller municipalities with higher proportions of workers in the agro-industry.

Another important factor identified in this study was immigration-related population composition, particularly the proportion of residents originating from countries with lower HDI classifications. Individuals’ gut microbiome and resistome are shaped by dietary habits, which are influenced by socio-cultural, and environmental factors (Wilson et al., 2020). Accordingly, people from different countries have different compositions of gut microbiota, as evidenced by global-scale analyses comparing sewage resistomes across regions (Hendriksen et al., 2019; Munk et al., 2022). Therefore, this parameter may contribute to the observed correlation between certain ARGs and immigration gradients (i.e., low-HDI), at least in part. However, human gut microbiomes evolve over time, and do not necessarily remain stable (Dwiyanto et al., 2021; Vangay et al., 2018). A study demonstrated that individuals lose gut microbiome diversity and function after migrating from non-Western countries to the United States, with the extent of change varying according to duration of residence (Dwiyanto et al., 2021; Vangay et al., 2018). In parallel, ARGs may be imported through acquisition during international travel, particularly to low-HDI countries (Arcilla et al., 2017; Ruppé et al., 2015). Many studies show that immigrants may have different preferences in travel destination, with returning to one’s country of origin (i.e., to visit friends and relatives) being especially common (Bacaner et al., 2004; Hung et al., 2013). Another study showed that gut microbiomes and resistomes can be disrupted during international travel, through acquisition of locally prevalent AMR (D’Souza et al., 2021). Our previous study analysing sewage resistomes across Calgary demonstrated similar findings, with wastewater AMR profiles across socio-demographically diverse neighborhoods exhibiting relative homogeneity during periods of minimal international travel in 2021 (owing to COVID-19-related travel restrictions), suggesting that continual introductions may contribute to observed variation (Lee et al., 2025). Our analysis does not enable differentiation between baseline microbiome differences and travel-related acquisition; these mechanisms may be confounded, with their relative contributions varying according to duration of residence and frequency and duration of international travel. More detailed individual-level data would be required to disentangle these effects. Improved integration of socio-demographic and exposure data may help clarify the primary mechanisms underlying immigration-associated AMR patterns.

### Distinct ARG profiles across socioeconomic and immigration gradients

By leveraging multivariate analysis followed by univariate correlation we identified that many ARGs were significantly associated with key socioeconomic determinants. Differential ARG profiles across gradients of low-HDI immigration and income suggest the existence of heterogeneous ecological and antimicrobial selection pressures operating across sub-populations. For instance, the relatively high abundance of tetracycline ARGs, particularly in cities with higher proportions of newcomers from low-HDI countries (e.g., Brooks, which has experienced substantial immigration from sub-Saharan African countries such as Sudan, Ethiopia, and Somalia over the past two decades) might be due to higher usage of tetracyclines in the countries from which corresponding sub-populations originated (StatsCan, 2021; Tebug et al., 2021). Indeed, tetracyclines have been widely used worldwide, particularly in livestock sectors, accounting for over 40% of total antibiotics used for food animals in the Global South (Gochez et al., 2022). In contrast, MLS ARGs were among genes negatively associated with low-HDI immigration, suggesting that certain ARGs may reflect resistance profiles associated with higher-income settings, where macrolide use is more prevalent (Klein et al., 2024). Furthermore, our findings confirmed the well-established negative association between AMR and income levels (Figs. 4 & 5). Notably, a greater diversity of ARGs (13 types versus 6) was negatively associated with income. This pattern may reflect broader and more complex selection pressures affecting economically disadvantaged populations, highlighting the need for more targeted and equitable healthcare investments.

### Limitations

There are several notable limitations of this work. Our surveillance did not include the 122 towns and villages serving small communities with <2000 individuals across Alberta, nor any populations residing in hamlets or low-connectivity rural areas. By focusing on populations served by large municipal wastewater treatment plants our results may underrepresent rural Alberta, including populations more closely associated with the agriculture and agri-food sectors. Nonetheless, our surveillance program captures a large and diverse segment of the population – representing approximately 40% of Alberta’s total population – thereby enabling characterization of population-level resistomes across urban-suburban interfaces, where population density is highest. While all communities in Alberta are supported by a single provincial health system (Alberta Health) which operates a single Antimicrobial Stewardship Program that is universally applied, we did not control for differences in local prescribing patterns. Most importantly, although our study incorporates a range of socio-economic and agro-industrial parameters grounded in conventional Canadian contexts, unmeasured factors may also contribute to population-level resistome patterns and explain their associations with societal variables, particularly at the individual level. Because our analyses were conducted at the municipal level, observed associations cannot be attributed to individuals or specific sub-populations. Establishing causality will require complementary study designs incorporating longitudinal individual-level data, targeted sub-population sampling, and experimental and/or quasi-experimental approaches.

### Societal determinants of AMR: One-Health and health equity perspectives

Our study focused on disentangling complex interconnections between AMR in humans, as reflected by wastewater-measured ARGs, and societal variables, leveraging an inclusive tool for monitoring population health. In this Canadian context, we show that associations between AMR and the human-animal interface may be overshadowed by broader societal factors, including income and immigration. However, this pattern likely reflects population-scale processes and may differ in certain localized environments, such as livestock feedlots or farms. Accordingly, more granular surveillance targeting specific sub-populations and exposure settings is warranted. Furthermore, our findings indicate that ARG burdens are not evenly distributed across populations but instead vary along socioeconomic and immigration gradients. These observations underscore the importance of incorporating equity-informed and context-specific strategies into AMR surveillance and intervention efforts.

## Ethics Statement

Ethics approval was obtained from the University of Calgary’s Conjoint Health Research Ethics Board (REB21-2025).

## Data Availability

Sequencing data were submitted to the NCBI Sequence Read Archive (SRA) under PRJNA1469283, and will be fully be published upon acceptance of this manuscript (https://dataview.ncbi.nlm.nih.gov/object/PRJNA1469283?reviewer=ng1lono4dmro4va1rkj37t84k7). Code and associated data analysis scripts are available via the first author’s GitHub repository (https://github.com/myjackson/PanAlberta-Project), and will be released as a packaged version, upon publication.

## Author Contributions

J.L. and M.D.P. conceptualized the study. J.L., C.G., E.A., N.A., and B.J.W. developed the methodology. N.A., B.J.W., and S.Bert. performed validation analyses. J.L. conducted the formal analysis. J.L., C.G., N.A., and S.Bert. curated the data. R.Z., T.A.M., and S.Bhat. provided resources. J.L. and M.D.P. prepared the original draft of the manuscript. All authors contributed to writing, review, and editing of the manuscript. M.D.P. and C.R.J.H. supervised the study. Project administration was performed by M.D.P., N.A., R.G.C., and C.R.J.H. Funding acquisition was performed by M.D.P., C.R.J.H., B.E.L., and X.P

## Acknowledgements

The authors are grateful for the contributions of the utility operators and management of Alberta’s municipal wastewater treatment plants.

