## Supplementary figures and tables for "Municipal wastewater surveillance reveals socioeconomic and immigration gradients in antimicrobial resistance across Alberta, Canada"

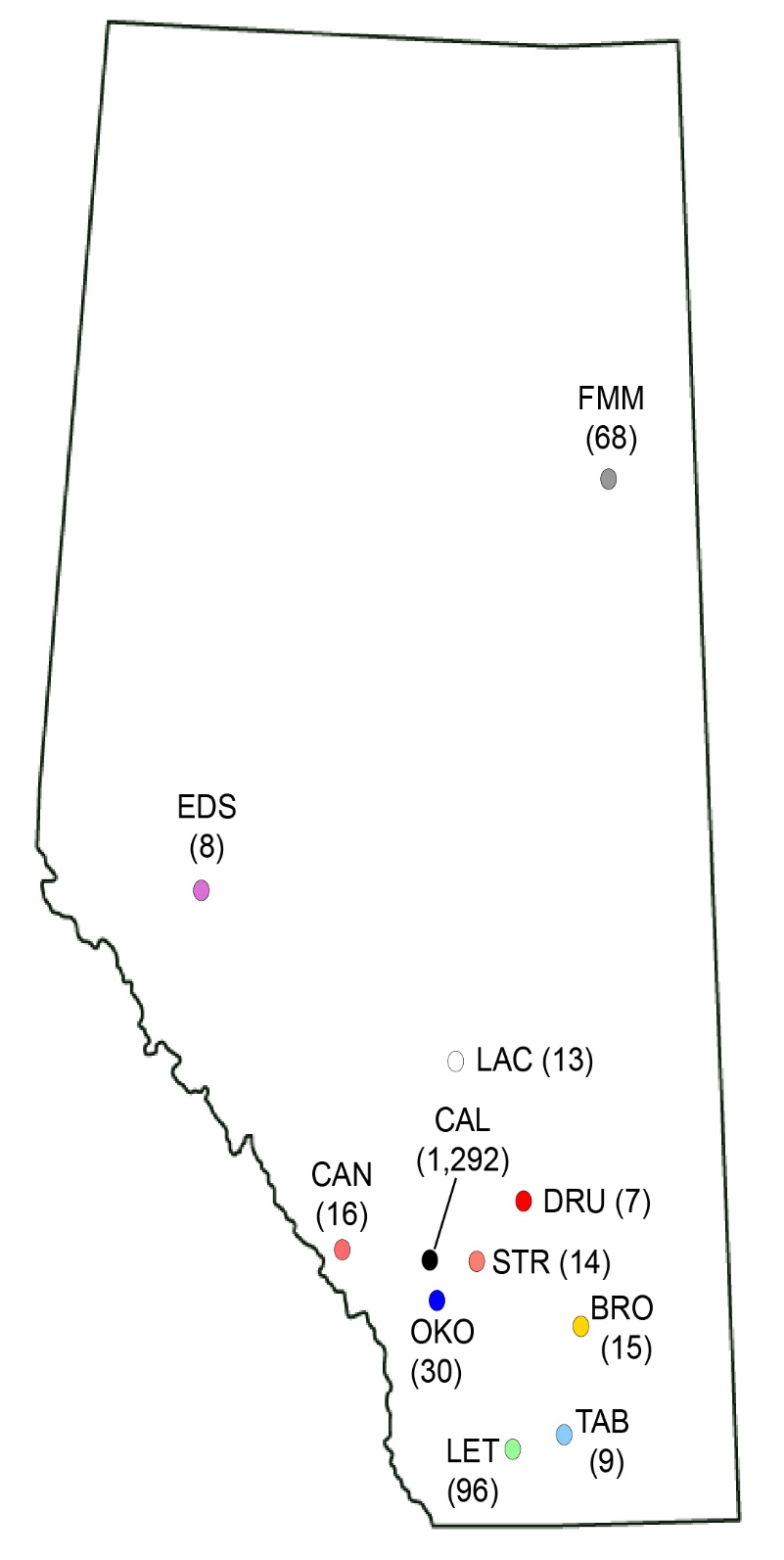

**Figure S1. Site map.** A total of 11 cities and communities across Alberta, Canada, were included, where population-level antimicrobial resistance genes were monitored through wastewater-based surveillance. The size of serving population for each wastewater treatment plant is provided in parentheses (unit: thousands of inhabitants). FMM=Fort McMurray, OKO=Okotoks, CAN=Canmore, CAL=Calgary, EDS=Edson, STR=Strathmore, LAC=Lacombe, BRO=Brooks, LET=Lethbridge, DRU=Drumheller, TAB=Taber.

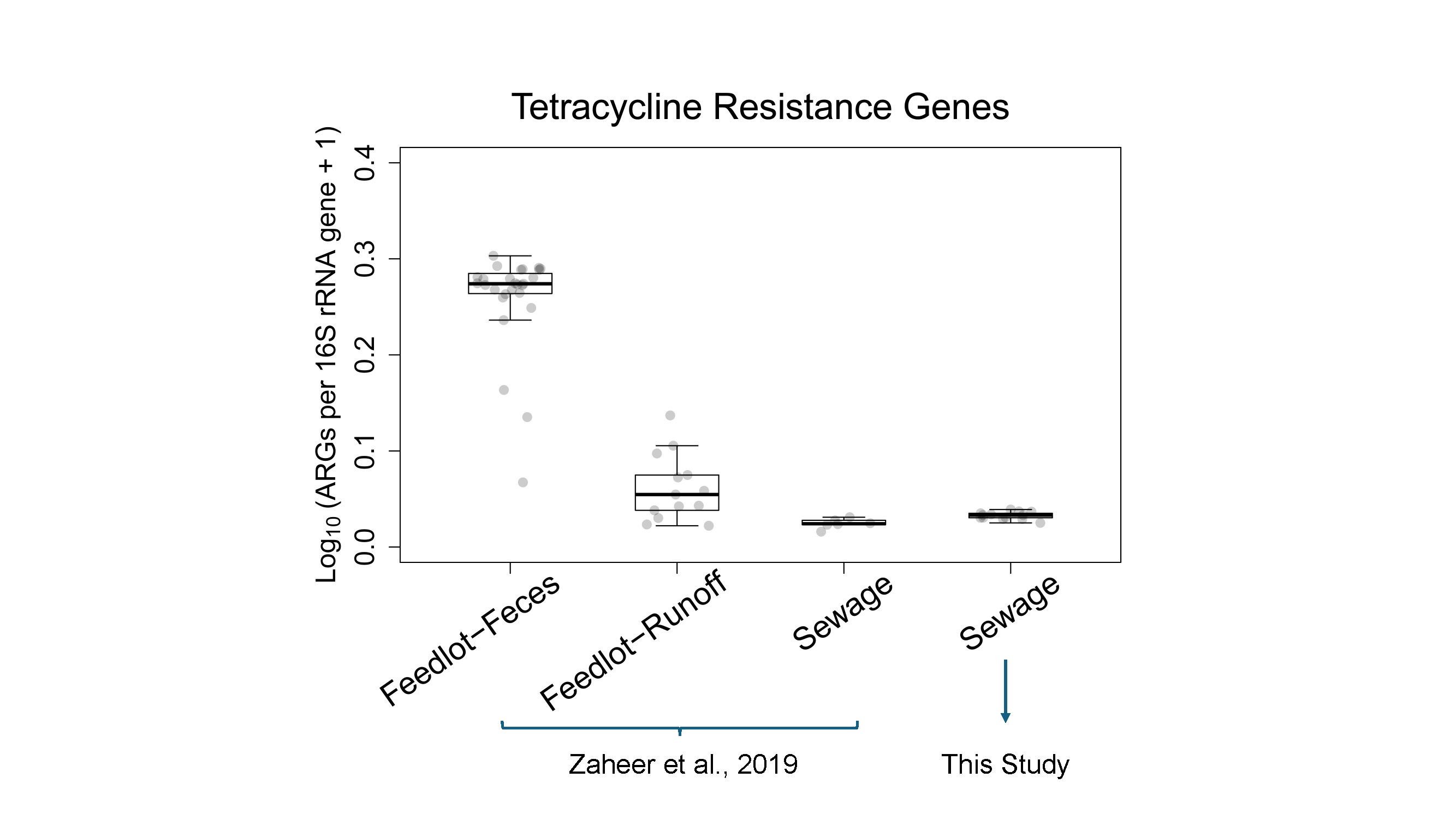
**Figure S2. Comparison of relative abundances of tetracycline resistance ARGs, occurring in cattle-related sources (i.e., feedlot cattle feces and runoff samples), and domestic sewage obtained from wastewater treatment plants, including one of our study sites (i.e., Calgary).** The data were sourced from Zaheer et al., 2019 or this study.

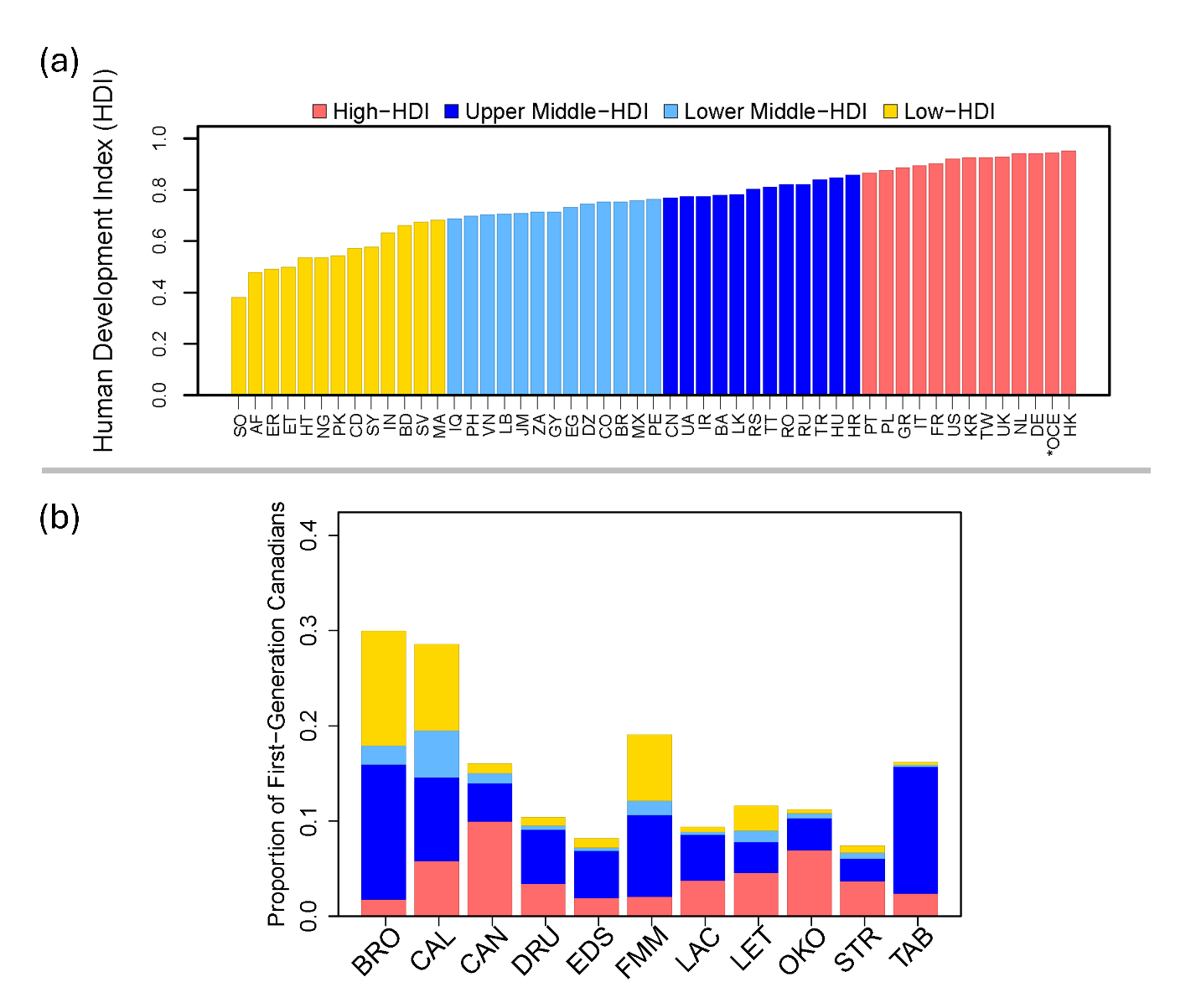
**Figure S3. Profiles of birth countries of first-generation Canadians across 11 sites.** (a) Human Development Index (HDI) of the countries of origin of first-generation Canadians. (b) Proportion of first-generation Canadians in the total population of each city, grouped by birth country and categorized into four HDI groups (high, upper-middle, lower-middle, and low).

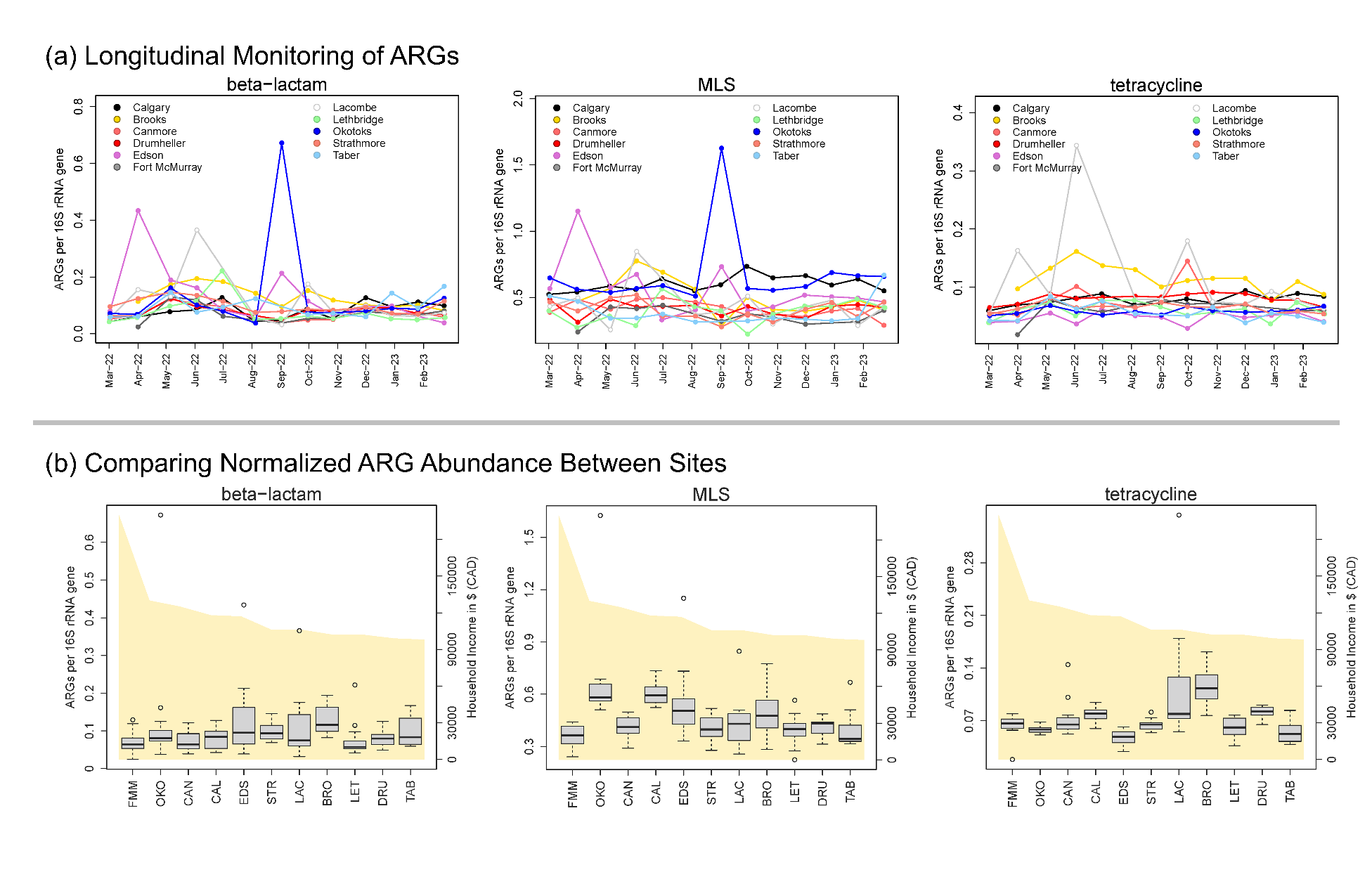

**Figure S4. Relative abundances of ARGs conferring resistance to three of the most frequently used antibiotics in the Canadian prairies over the course of monitoring (13 months during March-2022 – March-2023)****, exhibiting no visible temporal trends across sites.** (a) Longitudinal trends of ARG relative abundances. (b) Comparisons of ARG relative abundances across locations; the area plot (in yellow) represents annual household income ($CAD), uniquely associated with each site.

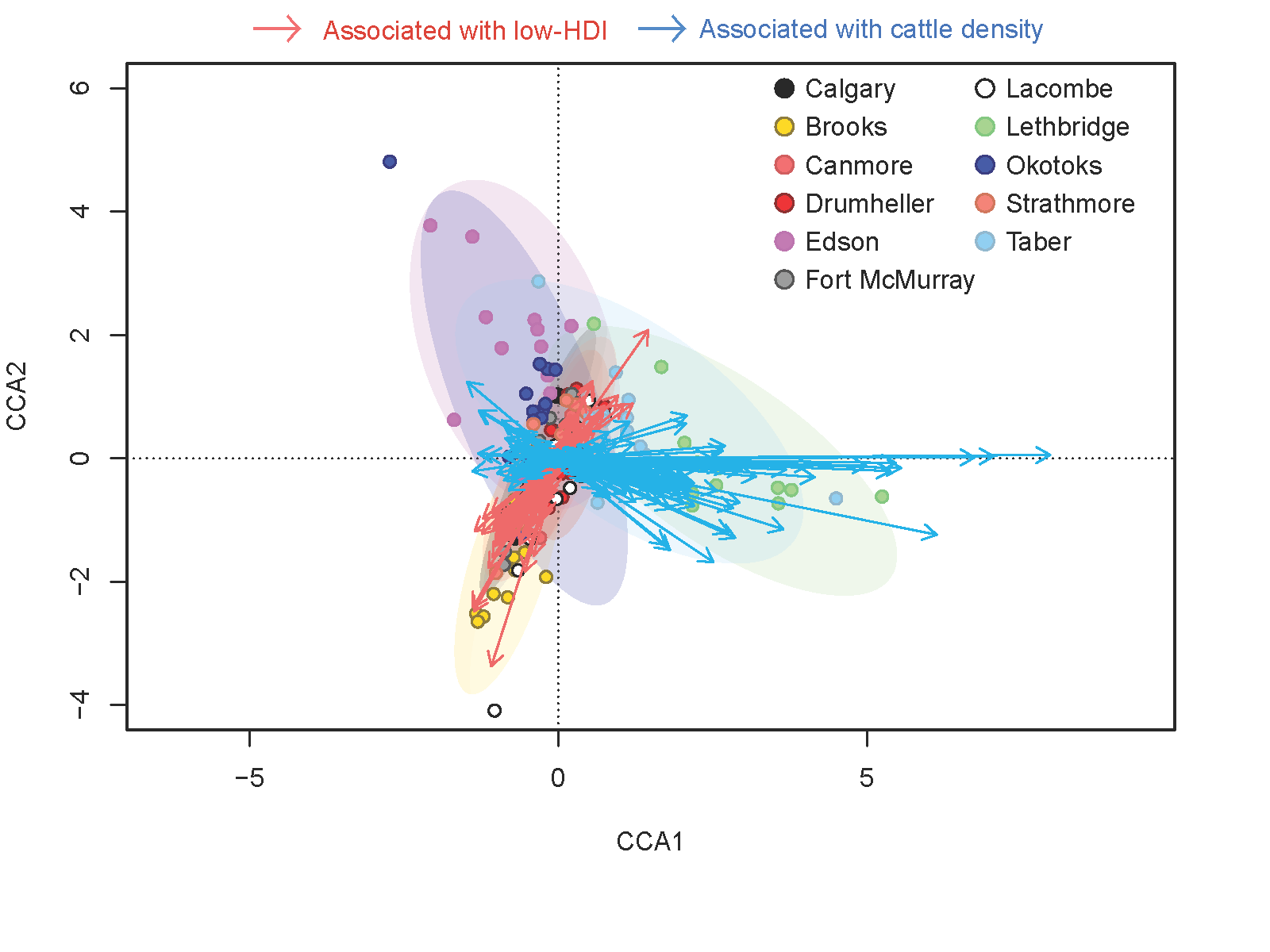

**Figure S5. The ARGs significantly correlated with two of the most significant societal variables, constraining CCA axis (i.e., immigration and cattle).** The genes significantly associated with L-HDI (see Fig. 2) were coloured in light red; the genes significantly associated with cattle (or income, but in an opposite direction) were coloured with light blue. A total of 200 ARGs, selected based on the residual sum of squares, were visualized, and further analysed for each group.

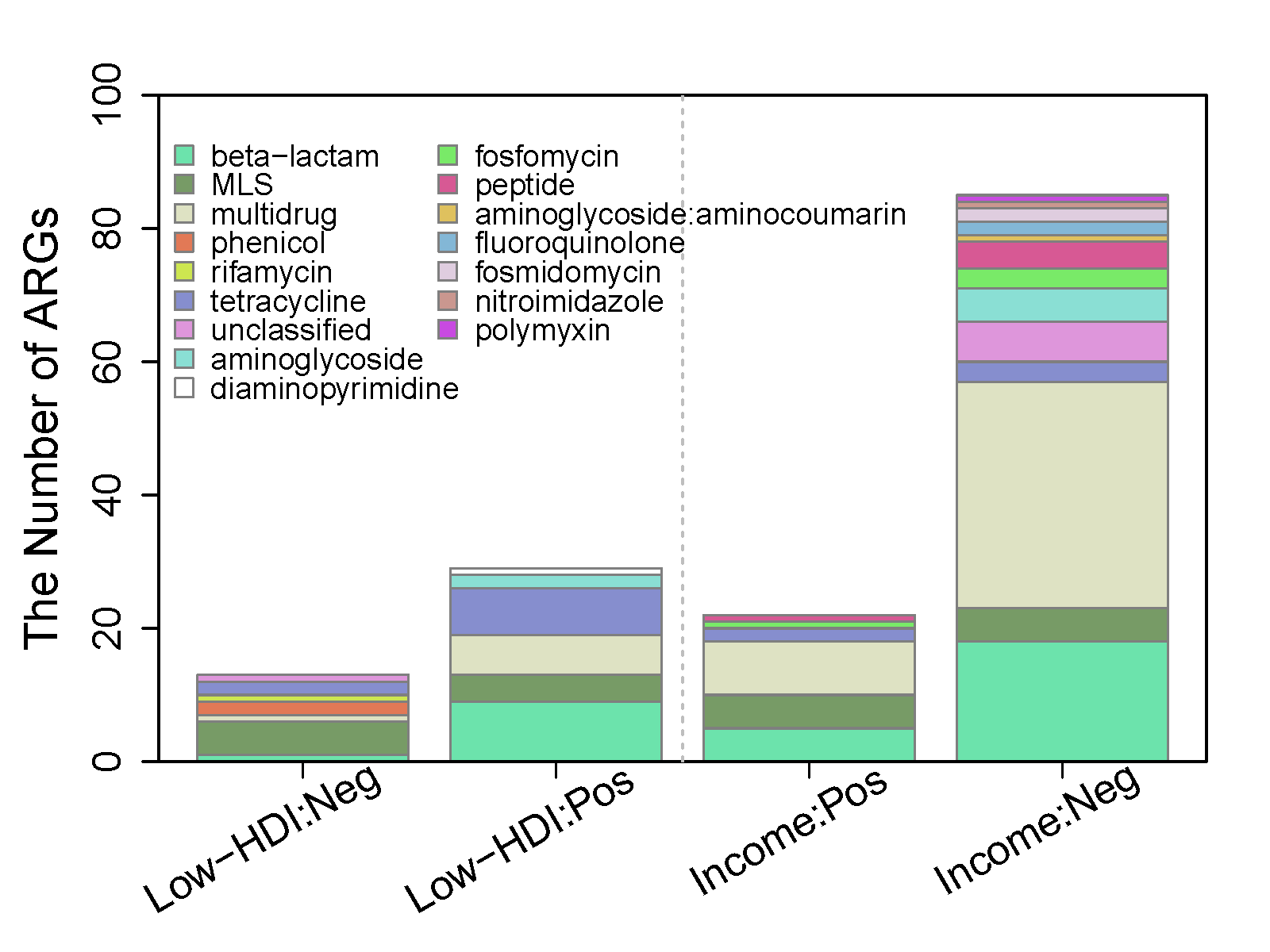

**Figure S6. The number of ARGs, occurred in four groups of key ARGs, associated significantly with immigration from Low-HDI or income (see Figs. 4 & 5).** Low-HDI:Neg and Low-HDI:Pos indicate key ARGs negatively or positively correlated with Low-HDI; Income:Pos and Income:Neg denote key ARGs positively or negatively correlated with income.
